# A blood-based miRNA signature with prognostic value for overall survival in advanced stage non-small cell lung cancer treated with immunotherapy

**DOI:** 10.1101/2021.10.31.21265722

**Authors:** Timothy Rajakumar, Rastislav Horos, Julia Jehn, Judith Schenz, Thomas Muley, Oana Pelea, Sarah Hofmann, Paul Kittner, Mustafa Kahraman, Marco Heuvelman, Tobias Sikosek, Jennifer Feufel, Jasmin Skottke, Dennis Nötzel, Franziska Hinkfoth, Kaja Tikk, Alberto Daniel-Moreno, Jessika Ceiler, Nathaniel Mercaldo, Florian Uhle, Sandra Uhle, Markus A Weigand, Mariam Elshiaty, Fabienne Lusky, Hannah Schindler, Quentin Ferry, Tatjana Sauka-Spengler, Qianxin Wu, Klaus F Rabe, Martin Reck, Michael Thomas, Petros Christopoulos, Bruno R Steinkraus

## Abstract

Immunotherapies have recently gained traction as highly effective therapies in a subset of late-stage cancers. Unfortunately, only a minority of patients experience the remarkable benefits of immunotherapies, whilst others fail to respond or even come to harm through immune related adverse events. For immunotherapies within the PD-1/PD-L1 inhibitor class, patient stratification is currently performed using tumor (tissue-based) PD-L1 expression. However, PD-L1 is an accurate predictor of response in only ∼30% of cases. There is pressing need for more accurate biomarkers for immunotherapy response prediction.

We sought to identify peripheral blood biomarkers, predictive of response to immunotherapies against lung cancer, based on whole blood microRNA profiling. Using three well characterized cohorts consisting of a total of 334 stage IV NSCLC patients, we have defined a 5 microRNA risk score (miRisk) that is predictive of overall survival following immunotherapy in training and independent validation (HR 2.40, 95% CI 1.37-4.19; *P* < 0.01) cohorts. We have traced the signature to a myeloid origin and performed miRNA target prediction to make a direct mechanistic link to the PD-L1 signalling pathway and PD-L1 itself. The miRisk score offers a potential blood-based companion diagnostic for immunotherapy that outperforms tissue-based PD-L1 staining.

## Introduction

Recently, immune checkpoint inhibitors (ICI), also known as immunotherapies, have emerged as a breakthrough treatment for advanced non-small-cell lung cancer (NSCLC) and other cancers^1^. These drugs target the inhibitory interaction between immune checkpoint receptors and their ligands, for example the Programmed-Death 1 (PD-1) pathway. While originally evolved to mediate self-tolerance, certain tumors have adapted to exploit these signalling pathways and thereby escape the immune response. If efficacious, immunotherapy augments T-cell immune responses against malignant cells and can deliver unprecedented clinical benefit. However, positive responses typically occur in only a minority of patients^2,3^. To date, five inhibitors of the PD-1/PD-L1 axis have been approved for advanced NSCLC: of these, pembrolizumab, nivolumab and cemiplimab target PD-1, whilst durvalumab and atezolizumab target its ligand PD-L1. Besides remarkable anti-tumor activity, these drugs are also associated with characteristic (and occasionally life-threatening) side effects, particularly immune-related adverse events and hyperprogression^4,5^. Taken together with the significant cost of these treatments, it is vital to administer immunotherapies only to those patients in whom the benefits are predicted to outweigh the risks.

The mainstay of efficacy prediction for PD-(L)1 inhibitors is the quantification of tumor PD-L1 expression, for which several FDA-approved commercial assays exist^6^. Present guidelines (ASCO and ESMO^7–9^) recommend first-line monotherapy with PD-(L)1 inhibitors for advanced NSCLC only when the PD-L1 tumor proportion scores (TPS) is at least 50%, that is ≥50% of viable tumor cells are positive for membrane staining of PD-L1. However, the necessity to biopsy and process tissue, spatial tumor heterogeneity, the use of different assay systems (three are FDA-approved) and different cut-offs, limit the accuracy and pose limitations to the consistency of this companion diagnostic approach^1,10,11^. Furthermore, because of a lack of alternative predictive biomarkers, potential responder populations outside the recommended PD-L1 TPS bracket are excluded from first-line PD-(L)1 inhibitor monotherapy. Accurately identifying responders independently of PD-L1 could expand the eligible population for immunomonotherapies, which would reduce the burden of chemotherapy toxicity for lung cancer patients.

Multiple lines of evidence suggest that information held in peripheral blood may be reflective of the immune response within the tumor microenvironment (TME). Associations have been found between immunotherapy response and peripheral blood counts^12^, peripherally expanded T-cell clones^13^, and monocyte subpopulations^14^. Based on this abundance of peripheral signal, we sought to define an immunotherapy response prediction signature from whole blood microRNA (miRNA) expression profiles. miRNAs are ∼22-nt short RNA molecules that regulate target messenger RNAs (mRNAs) through Watson-Crick base-pairing to miRNA response elements (MREs) frequently embedded in the 3’-untranslated region (UTR) of target genes^15^. This positions miRNAs as sophisticated regulators of gene expression, which can reflect immune status and activity^16^. To ensure a simple, robust, and reproducible platform, the IVD-certified PAXgene Blood RNA System (PAXgene) was used for the collection, lysis, and subsequent RNA stabilisation of whole blood samples to enable “pipetting-and cell sorting-free” sample collection in the clinic^17^. Analytical processes, including RNA extraction, library preparation, and next generation sequencing (NGS) were optimised to measure a whole blood, immune enriched, small RNA expression profile. Using several cohorts of stage IV NSCLC patients, we developed a model using miRNA expression profiles to predict overall survival (OS) following monotherapy with either pembrolizumab or nivolumab. We uncovered a myeloid enriched 5 miRNA-signature (miRisk score) which outperforms PD-L1 TPS and generalizes to an independent validation cohort. This miRisk score offers the prospect of a blood-based companion diagnostic for immunotherapy in advanced NSCLC.

## Results

### Clinical patient characteristics

A total of 334 patients were recruited from two sites and succesively assembled into three cohorts based on their time of recruitment and type of treatment; both training and validation cohorts received immunotherapy with either pembrolizumab or nivolumab, while the control ncohort received platinum doublet chemotherapy and pembrolizumab. Technical processing and measurement of samples was performed seperately for the training cohort and validation/control cohorts in order to maintain independence of cohorts and better estimate generalizable performance of predictive algorithms.

The 96 patients in the training cohort of stage IV NSCLC patients who received anti-PD-1 monotherapy comprised 77% (n = 74) pembrolizumab- and 23% (n = 22) nivolumab-treated patients; 58% adenocarcinomas (n = 56), 28% squamous cell carcinomas (n = 27), and 14% other NSCLCs (n = 13, including NSCLC, NOS and large-cell neuroendocrine lung carcinomas); 62.5% males; 91.7% former or current smokers (Table 1). Immunotherapy was administered in the first line to 49%, in the second line to 48%, and beyond the second-line to only 3% of the patients (Table 1). 71% had PD-L1 TPS ≥ 50%, 21% 1-49%, and 8% <1%, while ECOG performance status (PS) at the time of immunotherapy start was 0 in 37%, 1 in 58%, and 2 in 5% of cases. The clinical characteristics of the independent validation cohort are broadly similar and shown in Table 1, with notable significant differences in the distribution of immunotherapy substance and line of therapy. The independent control cohort consisted of 139 stage IV NSCLC patients treated with combined chemoimmunotherapy but who otherwise displayed broadly similar clincopathological features (Table 1).

**Table 1.** Clinical Characteristics of Patient Cohorts.

|  | Training | Independent Validation | p-value<sup>1</sup> | Control Cohort |
| --- | --- | --- | --- | --- |
| Treatment | Immunotherapy | Immunotherapy | Training vs. | Chemoimmunotherapy |
| Characteristic | (n=96) | (n=99) | Validation | (n=139) |
| Site |  |  | 0.0002 |  |
| Heidelberg | 96 | 84 |  | 139 |
| Grosshansdorf | - | 15 |  | - |
| Sex, no. (%) |  |  | 0.8708 |  |
| Male | 60 (62.5%) | 64 (64.6%) |  | 93 (66.9%) |
| Female | 36 (38.5%) | 35 (35.4%) |  | 46 (33.1%) |
| Age at enrollment, yr |  |  | 0.3820 |  |
| Mean ±SD | 67.6 ±9.4 | 66.4 ±9.4 | | 64.8 ±9.0 |
| Median (range) | 68.2 (38.9-86.7) | 66.4 (33.5-87.0) |  | 64.9 (37.6-84.8) |
| Histological Subtype, no. (%) |  |  | 0.6233 |  |
| Adenocarcinoma | 56 (58.3%) | 56 (56.6%) |  | 106 (76.3%) |
| Squamous cell carcinoma | 27 (28.1%) | 33 (33.3%) |  | 18 (12.9%) |
| Other | 13 (13.5%) | 10 (10.1%) |  | 15 (10.8%) |
| ECOG performance status, no. (%) |  |  | 0.3762 |  |
| 0 | 35 (36.5%) | 38 (38.4%) |  | 56 (40.3%) |
| 1 | 56 (58.3%) | 52 (52.5%) |  | 79 (56.8%) |
| 2 | 5 (5.2%) | 5 (5.1%) |  | 3 (2.2%) |
| 3 | - | 1 (1.0%) |  | 1 (0.7%) |
| NA |  | 3 (3.0%) |  |  |
| Smoking status, no. (%) |  |  | 0.5597 |  |
| Never | 8 (8.3%) | 5 (5.1%) |  | 12 (8.6%) |
| Former | 56 (58.3%) | 56 (56.6%) |  | 74 (53.2%) |
| Current | 32 (33.2%) | 38 (38.4%) |  | 53 (38.1%) |
| Therapy, no. (%) |  |  | 0.0025 |  |
| Nivolumab | 22 (22.9%) | 44 (44.4%) |  | - |
| Pembrolizumab | 74 (77.1%) | 55 (55.6%) |  | - |
| Platinum doublet + Pembrolizumab | - | - |  | 139 (100%) |
| Therapy Line, no. (%) |  |  | 0.0020 |  |
| 1 | 47 (49.0%) | 36 (36.4%) |  | 124 (89.2%) |
| 2 | 46 (47.9%) | 43 (43.4%) |  | 15 (10.8%) |
| 3 | 3 (3.1%) | 13 (13.1%) |  | - |
| >3 | - | 7 (7.1%) |  | - |
| PD-L1 TPS, no. % |  |  | 0.1018 |  |
| ≥50 | 68 (70.8%) | 56 (56.6%) | | 31 (22.3%) |
| 1-49 | 20 (20.8%) | 27 (27.3%) |  | 38 (27.3%) |
| <1 | 8 (8.3%) | 16 (16.2%) |  | 70 (50.4%) |

<sup>1</sup>a two-tailed t-test was performed for age, chi-squared test for all other variables

### Development of whole blood small RNA sequencing pipeline

Whole blood miRNA profiling is limited by the presence of a small number of highly abundant erythroid miRNAs, which limits the sequencing bandwidth available for the detection and accurate quantification of other RNA species. To mitigate this, we have used custom oligonucleotides in order to block miR-486-5p, miR-451a, and miR-16-5p during library preperation, which would otherwise occupy approximately 50% of the resulting reads (Fig.1a and Supplementary Fig. 1a-c). Paired sequencing of PAXgene samples using either an unblocked or blocked library preparation demonstrated a highly specific and efficient blocking of target miRNA species. The proportion of reads mapping to the three intended targets of blocking is efficiently reduced by 99.96% in blocked compared to unblocked libraries (Fig. 1c). The liberated sequencing bandwidth resulted in an increase of blocked reads per million (RPM) values, enabling a more accurate quantification of less abundant but potentially more informative immune cell derived miRNAs at a given sequencing depth (Fig.1d). The global miRNA expression profile was minimally affected as revealed by the high correlation coefficient (Fig. 1d, r = 0.99).

**Figure 1.**
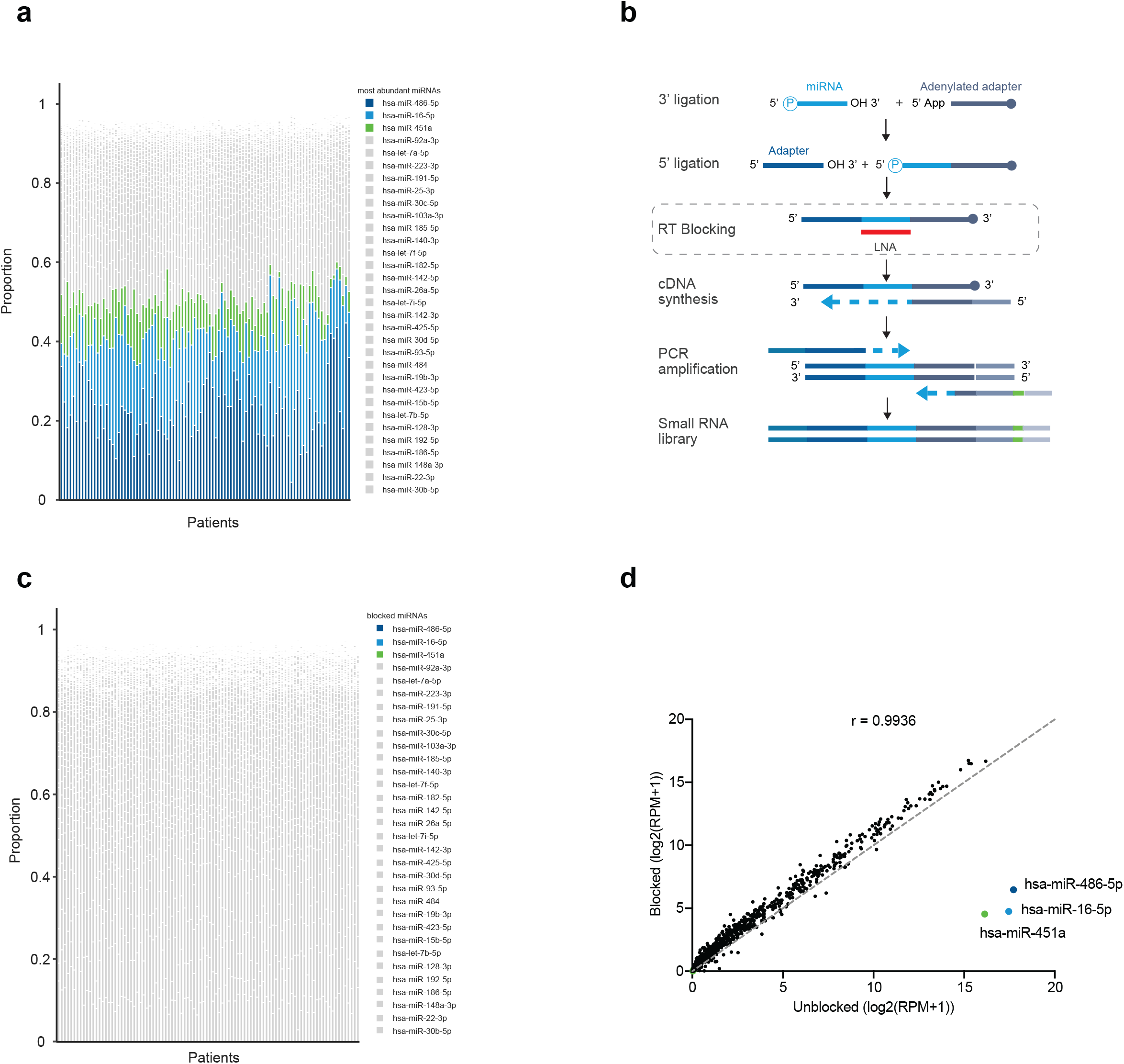
Whole blood small RNA sequencing pipeline with blocking of highly abundant miRNAs. **a** Sequencing of whole blood from 96 NSCLC patients revealed that ∼50% of reads per patient map to hsa-miR-486-5p, hsa-miR-16-5p, and hsa-miR-451a. **b** The standard miRNA library preparation protocol has been modified to block the incorporation of specific miRNAs using antisense locked nucleic acid (LNA) oligonucleotides that block the reverse transcription of target miRNAs. **c** The three blocking target miRNAs have been almost entirely eliminated from the sequencing libraries therefore increasing the sequencing bandwidth available for the detection of other miRNAs. **d** Mean miRNA expression following sequencing of unblocked or blocked libraries reveals specific depletion of the on-target miRNAs whilst maintaining a high correlation between th expression values of all other features (r = 0.99).

### Building a miRNA-based risk model for OS under PD-1 inhibitor monotherapy

miRNA expression profiles from patients in the training cohort (96 patients) were used to develop a model to predict OS of stage IV NSCLC patients receiving single agent immunotherapy of either pembrolizumab or nivolumab using a computational pipeline based on that described by Shukla et al.^18^. Models were optimised through a process of cross-validation to determine optimimal feature selelection parameters (Supplementary Fig. 2). Finally, this led to the generation of a linear predictor risk score: miRisk = (log(miR-2115-3p 26 RPM + 1) x 1.870) + (log(miR-218-5p RPM + 1) x 0.907) + (log(miR-224-5p RPM + 1) x 27 0.495) + (log(miR-4676-3p RPM + 1) x 1.309) + (log(miR-6503-5p RPM + 1) x 1.159), as 28 explained further in the Methods.

miRisk scores were calculated for all 96 patients in the training cohort (Table 1) and patients were separated into low/high risk groups based on the median risk score threshold (5.61). Patients in the low-risk group survive for significantly longer than those in the high-risk group (hazard ratio (HR) 3.98, 95% confidence interval (CI) 2.29-4.54; *P* < 0.01) (Fig. 2a). To quantify the generalizability of the model on a sample of independent patients, we applied the miRisk score to 99 patients in the validation cohort (Table 1), and stratified into low/high risk groups using the same cut-off (5.61). Patients in the low-risk group survived for significantly longer than those in the high-risk group (HR 2.40, 95% CI 1.37-4.19; *P* < 0.01) (Fig. 2b). The discriminative ability of the miRisk score was assessed using time dependent receiver operating characteristic (ROC) curve analysis to summarize OS at 6 months (Fig. 2g-i). The areas under the time dependent ROC curve (AUC) were 0.87 (95% CI 0.78-0.95) and 0.71 (95% CI 0.58-0.82) in the training and validation cohorts respectively, further supporting the relationship between the miRisk score and OS in stage IV NSCLC patients receiving immunotherapy at the clinically relevant timepoint of 6 months.

**Figure 2.**
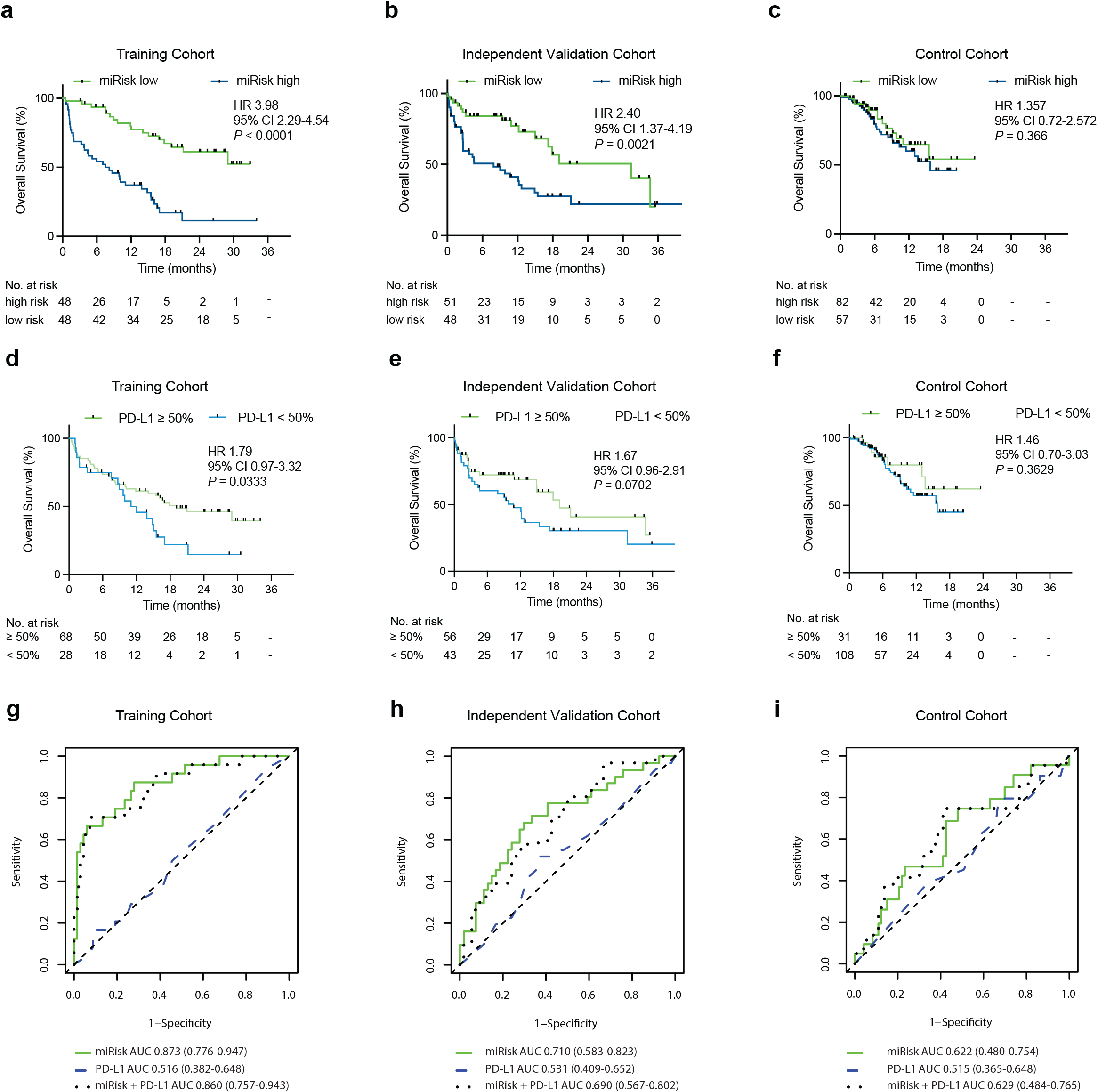
Overall survival of NSCLC patients stratified by miRisk score and PD-L1 TPS. **a-c** Comparison of OS between low/high miRisk score groups in the training (n=96), independent PD-(L)1 inhibitor monotherapy validation (n=99), and the chemoimmunotherapy control cohorts (n=139). Significant differences in OS are observed in the training and independent validation cohorts but not the control cohort. **d-e** Comparison of OS between patients stratified by PD-L1 TPS in the training (n=96), independent validation (n=99), and the control cohorts (n=139). The differences in OS only reach significance in the training cohort. Hazard ratios (HR) and 95% confidence intervals were calculated using a univariable Cox regression analysis; *P*-values were calculated using the log-rank test. All statistical analyses were two-sided. **g-h** Time dependent ROC curves (6 months) in the training, independent validation and control cohorts, as determined by the miRisk score, PD-L1 TPS or a model incorporating the miRisk miRNAs + PD-L1 TPS.

To explore whether non-linear models could reproduce and/or increase predictive performance, we trained and evaluated a random forest classifier on our dataset^19^. A random forest was fit to all miRNA expression features within the training cohort, without any prior feature selection, and the resultant median risk score was used to stratify patients into low/high risk groups. We observed significant differences in OS between the risk groups in both the training and independent validation cohorts (Supplementary Fig. 3a and 3b, 23 respectively; HR 4.94, 95% CI 2.82-8.66; *P* < 0.01; and HR 1.84, 95% CI 0.98-3.44; *P* = 0.03, respectively). Finally, permutation-based feature importance was used to identify the most informative miRNA features used by the random forest classifier. We observed 3/5 of the miRisk miRNAs, which were identified in the previously described approach, were amongst the most important features used by the random forest classifier including miR-1 2115-3p, the feature from the miRisk model with the greatest weighting (Supplementary Fig.2 3c).

The NGS measured expression levels of four of the five miRNAs that contribute to the miRisk score are observed to be significantly increased in high-risk patients (Fig. 3a-e). Only vmiR-218-5p is expressed at similar levels within the high- and low-risk groups (Fig. 3b). To technically validate our findings using an orthogonal method, we remeasured the miRNA expression profiles of 95 patients (one patient had insufficient RNA available) from the training cohort using qRT-PCR. This analysis recapitulated the NGS results for all five miRNAs with identical direction of change and similar degrees of significance (Fig. 3f-j).

**Figure 3.**
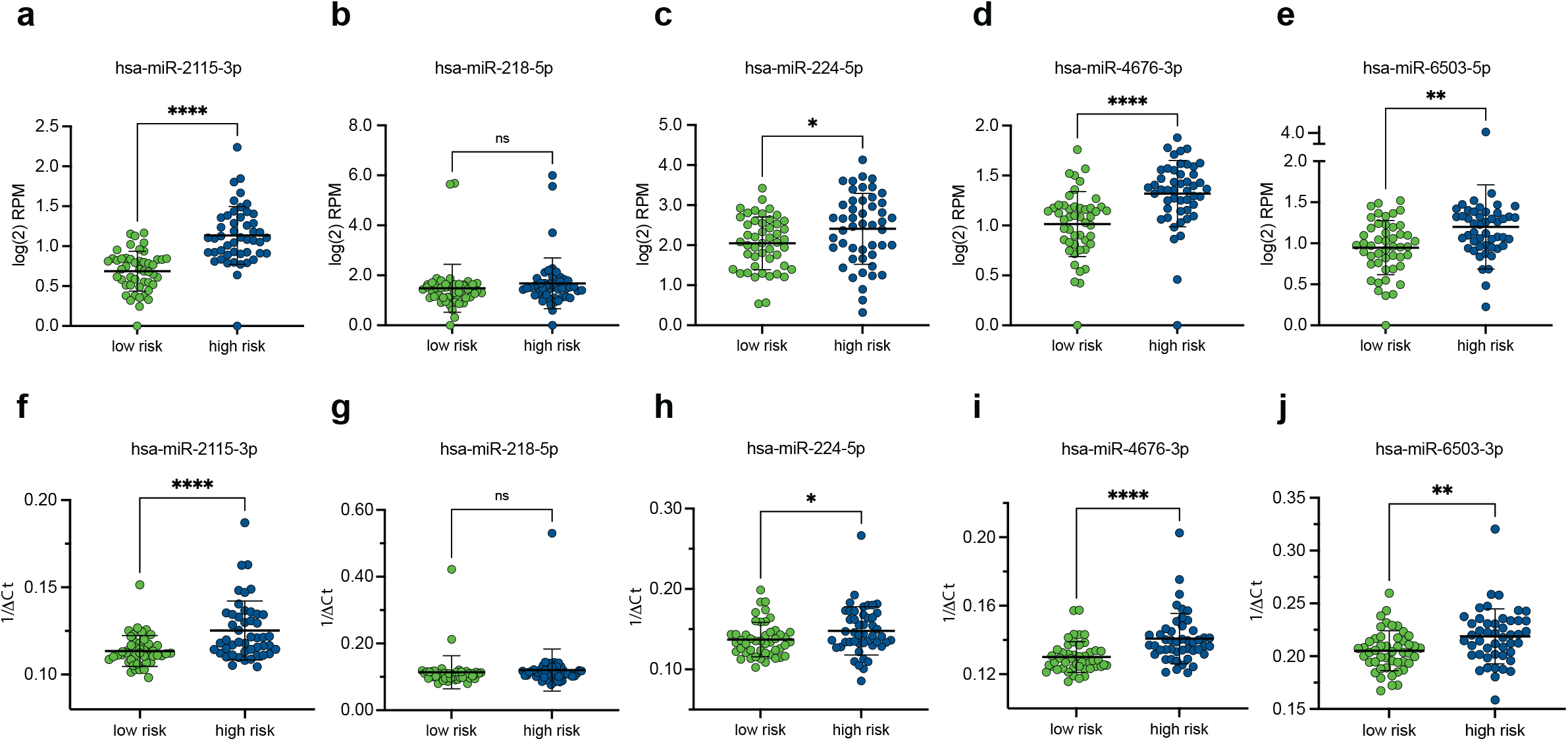
miRisk miRNA expression in low-risk and high-risk patients. **a-e** Relative expression levels of the 5 miRisk miRNAs between low-risk and high-risk patients measured by small RNA sequencing. **f-j** Relative expression levels of the 5 miRisk miRNAs between low-risk and high-risk patients measured by qRT-PCR. The mean expression of triplicate measurements is shown. qRT-PCR expression was normalized according to the ΔCt method (Ct_miRNA of interest_ - Ct_mean of HK miRNAs_). All statistical tests were two-tailed unpaired t tests. Error bars denote standard deviation. RPM = reads per million. * = *p*-value < 0.05, ** = *p*-value < 8 0.005, *** = *p*-value < 0.0005, **** = *p*-value < 0.00005, ns = not significant.

Together, we trained two entirely different models on our dataset and observed convergence on a subset of informative miRNA features. We technically validated the quantification of features by showing consistency between measurements from both NGS and qRT-PCR. Given the greater hazard ratio between risk groups predicted by the Cox based miRisk score compared to random forests, and its ease of interpretability, the miRisk score was chosen for further investigation.

### miRisk score is predictive for survival following PD-1 inhibitor monotherapy in real world clinical cohorts

The miRisk based stratification of late stage NSCLC patients receiving immunotherapy may be a result of miRisk correlation to either the treatment efficacy (predictive), or the patient outcome independent of treatment (prognostic). This would ideally be investigated in a control cohort who were not treated with immunotherapy. Unfortunately, such patients were not available, because current guidelines mandate use of PD-(L)1 inhibitors in the first-line treatment of all newly diagnosed stage IV NSCLC patients without actionable genetic alterations^7–9^. Therefore, instead we used as a control cohort patients treated with combined chemoimmunotherapy, given previous reports that immunotherapy specific biomarkers fail to predict survival outcomes in these patients (Table 1)^20–22^.

Following miRisk stratification of patients in the control cohort we observed no significant difference in survival between those in the low-risk versus high-risk groups (HR 1.36, 95% CI 0.72-2.57; *P* = 0.37) (Fig. 2c). Similarly, we observed no significant prediction of 6 month survival with the miRisk score in the control cohort using the time dependent ROC analysis (Fig. 2i). The interpretation of these results is, however, limited by the potential mixed effects of concurrent treatment with immunotherapy and chemotherapy, and the shorter duration of patient follow up relative to the other study cohorts. Still, in these real world clinical cohorts, these results provide evidence of miRisk as a predictor of survival following immunotherapy monotherapy.

### Evaluating the miRisk score in a clinical context

To explore the association between the miRisk score and other clinical covariates with respect to OS, both univariable and multivariable Cox regression were performed within all three clincal cohorts (Table 2). In addition to the miRisk score, we included clinicopathological covariates of relevance to immunotherapy response in these models. Based on the univariable analysis, we observed significant associations between OS and miRisk in both the training (HR 4.35, 95% CI 2.41-7.85; *P* < 0.01) and validation (HR 2.47, 22 95% CI 1.36-4.48; *P* < 0.01) cohorts but not the control (HR 1.36, 95% CI 0.70-2.64; *P* = 0.37) cohort, in concordance with previous analyses. Similarly, a trend is observed between a higher PD-L1 TPS and longer OS in the training and validation cohorts with greater significance than that observed in the control cohort. An association between ECOG and OS is observed only in the validation cohort (HR 2.82, 95% CI 1.47-5.41; *P* < 0.01). Across all three cohorts correlations between blood counts and OS are observed, with significant association to absolute neutrophil count (ANC), and non-significant associations to absolute lymphocyte count (ALC). The beneficial effect from both low ANC and high ALC is consistent with a previously published predictive signature derived from peripheral blood counts^12^, however the observed association in the control cohort may be suggestive of a prognostic element to this biomarker.

**Table 2.** Univariable and multivariable Cox regression analysis of miRisk and clinical covariates.

|  |  |  |  |  |  |  |
| --- | --- | --- | --- | --- | --- | --- |
| Training Cohort |  |  |  |  |  |  |
| Overall survival | Univariable Analysis |  |  | Multivariable Analysis |  |  |
| Covariate | HR | 95% CI | p-value | HR | 95% CI | p-value |
| PD-L1 (≥50%) | 1.81 | 1.04-3.16 | 0.036 | 1.48 | 0.69-3.17 | 0.314 |
| ECOG<sup>1</sup> (>0) | 1.49 | 0.84-2.64 | 0.175 | 1.41 | 0.75-2.66 | 0.284 |
| Gender (male vs female) | 0.82 | 0.48-1.40 | 0.457 | 0.86 | 0.48-1.56 | 0.623 |
| Age (>75 years) | 1.27 | 0.67-2.41 | 0.470 | 1.25 | 0.58-2.69 | 0.577 |
| Therapy line incl. <IV (>2) | 1.43 | 0.35-5.93 | 0.621 | 1.61 | 0.34-7.58 | 0.549 |
| Substance (Pembro vs Nivo) | 0.60 | 0.33-1.07 | 0.085 | 0.96 | 0.44-2.10 | 0.926 |
| Histology (Non-adeno vs adeno) | 1.19 | 0.70-2.04 | 0.519 | 1.22 | 0.68-2.19 | 0.513 |
| Smoking status (ever smoker vs other) | 0.67 | 0.29-1.57 | 0.356 | 0.68 | 0.24-1.94 | 0.472 |
| ANC<sup>2</sup> (>7.5) | 1.87 | 1.09-3.22 | 0.023 | 1.24 | 0.68-2.27 | 0.485 |
| ALC<sup>3</sup> (>1) | 0.61 | 0.35-1.04 | 0.068 | 0.88 | 0.47-1.65 | 0.683 |
| miRisk (high vs low) | 4.35 | 2.41-7.85 | <0.001 | 3.83 | 1.99-7.37 | <0.001 |
| Independent Validation Cohort |  |  |  |  |  |  |
| Overall survival | Univariable Analysis |  |  | Multivariable Analysis |  |  |
| Covariate | HR | 95% CI | p-value | HR | 95% CI | p-value |
| PD-L1 (≥50%) | 1.67 | 0.95-2.94 | 0.073 | 1.54 | 0.45-5.31 | 0.492 |
| ECOG<sup>1</sup> (>0) | 2.82 | 1.47-5.41 | 0.002 | 4.46 | 2.15-9.24 | <0.001 |
| Gender (male vs female) | 1.07 | 0.6-1.93 | 0.816 | 0.81 | 0.43-1.52 | 0.508 |
| Age (>75 years) | 1.66 | 0.85-3.24 | 0.141 | 1.81 | 0.87-3.81 | 0.115 |
| Therapy line incl. <IV (>2) | 1.36 | 0.73-2.54 | 0.327 | 1.32 | 0.67-2.58 | 0.421 |
| Substance (Pembro vs Nivo) | 0.68 | 0.39-1.19 | 0.176 | 1.18 | 0.35-4.01 | 0.789 |
| Histology (Non-adeno vs adeno) | 1.63 | 0.93-2.85 | 0.088 | 1.59 | 0.81-3.12 | 0.174 |
| Smoking status (ever smoker vs other) | 1.92 | 0.46-7.96 | 0.367 | 4.53 | 0.98-20.99 | 0.054 |
| ANC<sup>2</sup> (>7.5) | 1.98 | 1.08-3.62 | 0.027 | 1.06 | 0.54-2.11 | 0.862 |
| ALC<sup>3</sup> (>1) | 0.57 | 0.32-0.99 | 0.047 | 0.68 | 0.38-1.22 | 0.200 |
| miRisk (high vs low) | 2.47 | 1.36-4.48 | 0.003 | 2.49 | 1.16-5.34 | 0.019 |
| Control Cohort |  |  |  |  |  |  |
| Overall survival | Univariable Analysis |  |  | Multivariable Analysis |  |  |
| Covariate | HR | 95% CI | p-value | HR | 95% CI | p-value |
| PD-L1 (≥50%) | 1.46 | 0.64-3.32 | 0.364 | 1.59 | 0.65-3.87 | 0.307 |
| ECOG<sup>1</sup> (>0) | 1.66 | 0.85-3.25 | 0.136 | 2.13 | 1.05-4.35 | 0.037 |
| Gender (male vs female) | 1.14 | 0.59-2.20 | 0.698 | 1.06 | 0.52-2.16 | 0.878 |
| Age (>75 years) | 0.75 | 0.27-2.12 | 0.590 | 0.52 | 0.17-1.56 | 0.241 |
| Histology (Non-adeno vs adeno) | 0.93 | 0.42-2.03 | 0.851 | 0.69 | 0.31-1.53 | 0.360 |
| Smoking status (ever smoker vs other) | 0.90 | 0.28-2.91 | 0.854 | 0.76 | 0.21-2.68 | 0.664 |
| ANC<sup>2</sup> (>7.5) | 0.47 | 0.25-0.90 | 0.022 | 0.37 | 0.18-0.75 | 0.006 |
| ALC<sup>3</sup> (>1) | 0.96 | 0.50-1.82 | 0.893 | 0.91 | 0.45-1.82 | 0.787 |
| miRisk (high vs low) | 1.36 | 0.70-2.64 | 0.369 | 2.52 | 1.20-5.30 | 0.015 |

<sup>1</sup>Eastern Cooperative Oncology Group (ECOG) performance status; <sup>2</sup>absolute neutrophil count; <sup>3</sup>absolute lymphocyte count

Based on the multivariable analysis (Table 2), the miRisk score is an independent significant predictor of OS in all three cohorts. Additionally, ECOG is associated with OS in the validation and control cohorts. The association between blood counts and OS is attenuated when controlling for other covariates in the training and validation cohorts, but not the control cohort where ANC remains an independent significant predictor (HR 0.37, 95% CI 0.18-0.75; *P* < 0.01). Together, these analyses would suggest that the miRisk score may have a prognostic association across the three studied cohorts. This may be indicative of the general prognosis of late stage NSCLC or rather related to the control cohort having also received immunotherapy as part of their treatment regime, and an association to the efficacy of this part of the treatment.

### miRisk score outperforms PD-L1 histology as a predictive biomarker in advanced NSCLC

To test the performance of the established gold standard biomarker in our clinical samples, we stratified patients within the training, validation and control cohorts based on PD-L1 TPS (≥50% vs. <50%). There is a significant association between OS and PD-L1 TPS in the training cohort and a non-significant trend within the validation cohort (Fig. 2d-e). No association is noted within the control cohort (Fig. 2f). The magnitude of the hazard ratio associated with a low PD-L1 TPS is less than that associated with a high miRisk score in both training and validation cohorts. To further quantify the increase in predictive performance of the miRisk score over PD-L1 TPS, we calculated the difference in time dependent ROC AUC at 6 months using either the miRisk score or PD-L1 TPS over 1000 bootstrapped datasets. ROC AUCs increased by 0.36 (95% CI 0.22-0.50) and 0.18 (95% CI 0.01-0.35) in the training and validation datasets respectively, when classified by miRisk score compared to PD-L1 TPS (Fig. 2g-h). To explore any potential additive performance, we retrained a Cox model using PD-L1 TPS in addition to the five previously identified signature miRNAs (miRisk+PD-L1). Using this model, we observed a difference in time 6 dependent ROC AUC of -0.01 (95% CI -0.04-0.02) and 0.01 (95% CI -0.04-0.05) in the training and validation cohorts respectively, demonstrating no significant incremental improvement in the miRisk + PD-L1 score performance compared to the miRisk score (Fig. 9 2g-h).

Together, this provides evidence of significantly superior performance of the blood-based miRisk score compared to tissue-based PD-L1 TPS when predicting OS at 6 months. These results corroborate the limitations of PD-L1 TPS as an immunotherapy efficacy predictor and are consistent with a recent meta-analysis that found PD-L1 TPS to be predictive of immunotherapy response in only approximately 30% of patients across a range of tumour types^23^.

### Predictive miRNAs are of myeloid origin

To deconstruct the contribution of distinct blood cell types to the mixed-cell PAXgene whole blood profile, we sequenced small RNAs of 10 purified cell populations sorted to high purity (Supplementary Data 2) from blood donated by 12 healthy volunteers between 50 and 60 years old (Supplementary Fig. 4). This enabled the creation of a blood cell type miRNA expression atlas and an exploration into the cell of origin of the 5 miRisk miRNAs. To integrate the blood cell type miRNA expression atlas with PAXgene whole blood miRNA expression profiles, a custom scaling factor was applied to the individual miRNA expression data, based on cell type abundance and intracellular RNA content (see Methods for calculation and Supplementary Data 3). This enabled a deconvolved picture of PAXgene miRNA expression based on the relative contributions of the dominant peripheral blood cell types (Fig. 4a). The dominance of RBCs in PAXgene small RNA preparations is apparent, as observed by 259 whole blood miRNAs predominantly originating from RBCs. The next most dominant cell type in terms of contribution to the PAXgene profile are platelets, which are the major source of a further 109 miRNAs. miRNAs that show immune cell restricted expression are in the minority (Fig. 4a).

**Figure 4.**
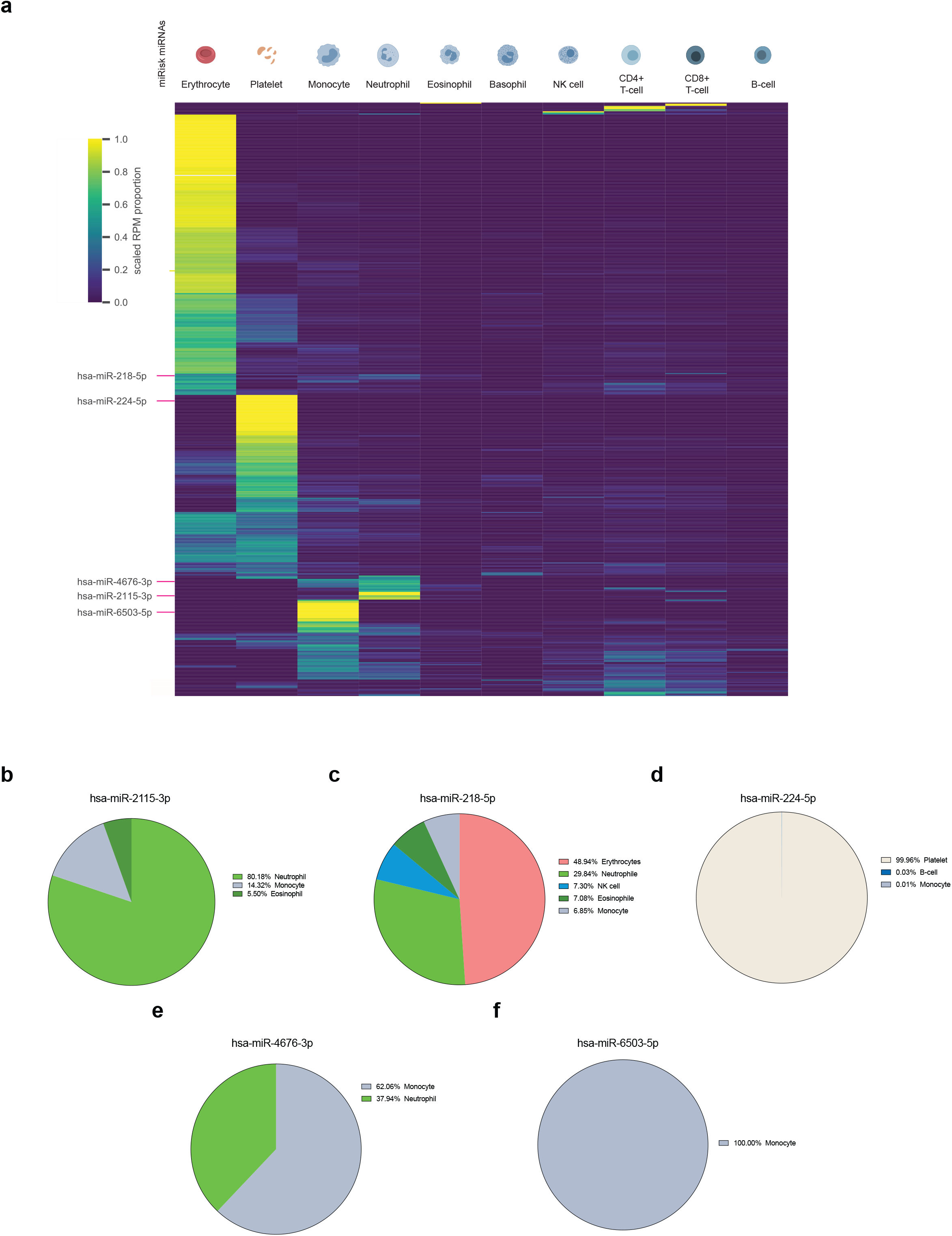
Cellular origin of miRisk miRNAs in peripheral whole blood. **a** Relative distribution of PAXgene detected miRNAs across 10 purified cell types. The 5 miRisk miRNAs are indicated. **b-f** The relative contributions and cellular origin of the 5 miRisk miRNAs. miRNA RPM values were scaled by cell type specific small RNA content per cell and abundance in blood. Per miRNA the scaled values of all measured cell types were indexed to 100%. RPM = reads per million.

Despite the majority of PAXgene detected miRNAs originating from RBCs, we observed a striking cell type specific expression pattern miRisk miRNAs in myeloid cells (miR-2115-3p, miR-4676-3p, and miR-6503-5p), and platelets (miR-224-5p) (Fig. 4b-f). miR-2115-3p, which contributes most highly to the weighted miRisk score, is exclusively expressed in the myeloid cell types of neutrophils, eosinophils and monocytes (Fig. 4b). Within the training cohort we observed a strong positive correlation between miR-2115-3p expression and the ANC, but not the eosinophil or monocyte counts (Supplementary Fig. 5). In contrast, miR-6503-5p is exclusively expressed in monocytes yet is uncorrelated to the monocyte count (Supplementary Fig. 5b). This would suggest that miR-6503-5p expression per cell is lower within peripheral monocytes of responder patients. Other miRisk miRNAs display weaker but significant correlations to their cell of origin (Supplementary Fig. 5) and thus may contribute to the risk profile as both a measure of peripheral cell counts and/or immune phenotype.

### Pathway analysis of predictive miRNAs and interaction with PD-1/PD-L1 signalling

To further characterize the mechanistic link between the miRisk miRNAs and immunotherapy response, we performed bioinformatic target prediction and pathway analysis. Employing the TargetScan algorithm^24^, we identified predicted target genes for both the 44 miRNAs significantly associated with OS (Supplementary Data 1), and the 5 miRNAs that contribute to the miRisk score. These gene sets were then used as inputs for pathway enrichment analysis using the KEGG database^25^. PD-L1 pathway genes are significantly enriched amongst targets of the 44 miRNA shortlist (1.22-fold enrichment, adjusted *P* = 4.8×10^−5^) and display a trend towards enrichment within the targets of the miRisk miRNAs (1.22-fold enrichment, adjusted *P* = 0.05) (Supplementary Data 4, Supplementary Fig. 6). Interestingly, this analysis revealed predicted direct interactions between PD-L1 (CD274) itself and the miRisk miRNAs miR-2115-3p, miR-4676-3p and miR-6503-5p (Supplementary Fig. 7a). Furthermore, the MAPK signalling pathway was identified as a highly enriched pathway amongst target genes of the miRisk miRNAs (1.21-fold enrichment, adjusted p-value = 1.6×10^−3^) (Supplementary Fig. 6b). This further suggests a potential functional link as the PD-L1/PD-1 pathway is upregulated by upstream signalling through the MAPK pathway^26^.

## Discussion

Response to immunotherapy is likely governed by the complex interplay between tumor and immune dependent factors, inherently limiting response prediction based on single biomarkers (e.g. PD-L1) or unilateral tumor-derived parameters (e.g. plasma tumor mutational burden (pTMB)). Instead, integrating multiple omics covariates may offer a better reflection of this complexity and thus a more accurate evaluation of risk and benefit^27–30^. Here we describe the discovery, validation and mechanistic insight into a 5 miRNA risk score (miRisk) measured in peripheral blood that predicts OS of stage IV NSCLC patients receiving PD-1 inhibitor monotherapy, and which performs better than the current standard of care, tissue PD-L1 TPS. We anticipate that as the evidence-based algorithms of clinical practice continue to evolve, such companion or complementary diagnostics will be vital to further refine patient management and deliver on the promise of personalized and precision medicine.

In this study we used peripheral blood miRNA profiles as a proxy for phenotypic inference into the tumor immune microenvironment (TME) and its susceptibility to immunotherapy. Our miRisk score likely captures and integrates information from multiple peripheral immune sources, reflective of the systemic immune status. Deconvolution of the PAXgene small RNA profiles into individual cell types revealed a remarkable myeloid specific expression pattern, largely originating from monocytes and neutrophils (miR-2115-3p, miR-4676-3p, miR-6503-5p), and platelets (miR-224-5p). Only miR-218-3p showed a heterogeneous pattern of expression (Fig.4c). Our findings are consistent with the growing evidence that peripheral biomarkers are predictive of response to immunotherapy^12–14,31,32^ including previous reports of miRNAs^33–36^. Myeloid cells, particularly monocytes but also neutrophils, have become increasingly recognized key players configuring the TME^37–39^. Myeloid Derived Suppressor Cells (MDSCs) have been shown to play critical roles in establishing an immunosuppressive TME and their subsequent response to immune checkpoint blockade^40,41^. Furthermore, platelets and more specifically tumor educated platelets have been described to undergo transcriptional changes upon transit through the TME and maintain memory of these events when they return to the periphery^42–44^. Given this body of evidence, and our observed correlation between miRisk miRNAs and peripheral blood cell counts (particularly miR-2115-3p and neutrophils), it is likely that some of the miRisk performance is derived from an indirect measure of blood count. However, the demonstration of the miRisk score as an independent significant predictor of OS in multivariable Cox analysis (Table 2) is evidence that it captures additional information of relevance to the efficacy of immunotherapies. Our work also provides insight into the mechanistic validity of the miRisk score. We have used bioinformatic analyses to reveal an overrepresentation of miRisk miRNA targets within the PD-1/PD-L1 signaling pathway, including multiple predicted direct interactions with PD-L1 itself Together we propose a possible mechanism whereby lower expression of the miRisk miRNAs in responder patients may correspond to a derepression of signaling through the PD-1/PD-L1 pathway in peripheral immune cells which is maintained upon their migration into the TME (Supplementary Fig. 7b). This could establish a TME in which immunosuppression depends more heavily on the PD-1/PD-L1 axis, and therefore could also be particularly susceptible to pharmacological inhibition of this pathway.

A reliable blood-based companion diagnostic biomarker for immunotherapy in NSCLC has the potential to dramatically improve patient care and outcomes. Some examples of potential clinical applications include the selection of suitable patients with PD-L1 <50% for PD-1 inhibitor monotherapy, selection of suitable PD-L1-high patients for immunomonotherapy *vs*. chemoimmunotherapy, selection of IO-pretreated patients in later lines for re-exposition to PD-(L)1 inhibitor therapy, and the decision of whether to continue PD-(L)1 inhibitor therapy beyond 2 years in patients with ongoing responses. In addition, due to the mechanistic underpinnings, serial miRisk measurements might provide a practical, non-invasive aid for translational studies aiming to characterize the temporal dynamics and immunobiology of tumor escape from immunotherapy in NSCLC, for example by helping identify suitable time-points for tissue rebiopsies under immunotherapy.

Specific methodological strengths of our study are the custom blocking of highly abundant erythroid miRNAs, which facilitated the study of less frequent, but immunologically more relevant miRNA species; the comprehensive bioinformatic evaluation including consistency of the results from two independent analyses based on Cox and random forest models; as well as the utilization of well-sized independent patient cohorts treated in two German lung cancer centers of excellence, with deep clinical annotation and long standing expertise in prospective biobanking^45^. At the same time, we acknowledge several limitations of the present work. To date, the miRisk score as a predictor of patient OS on immunotherapy has only been validated in a single independent patient cohort. We acknowledge that the predictive versus prognostic nature of the miRisk score has not been definitively addressed. This was hindered through a suboptimal control cohort which also received immunotherapy (in combination with chemotherapy). Furthermore, the shorter period of follow up for patients within the control cohort may have skewed comparisons to the training and validation cohorts. Until the potential predictive power of the miRisk score to immunotherapy response is further delineated it is unable to be applied to clinical therapeutic decision making for late stage NSCLC. We are currently recruiting additional NSCLC patients to further assess the generalized performance of the model in further validation cohorts and control cohorts that have been treated with chemotherapy alone. Additionally, although we propose a systemic mechanism which could in principle be independent of primary tumor site, we have not yet assessed the miRisk score in cancers other than NSCLC. We are currently collecting additional patients suffering from other cancer types to evaluate whether miRisk is universally applicable. Lastly, the hypothetical mechanism behind miRisk is largely based on bioinformatic analyses at present. Future work will be needed to demonstrate the interaction between miRisk miRNAs and PD-L1 expression directly, both in circulating blood cells and the TME.

In summary our findings demonstrate the potential utility of peripheral blood-based miRNAs as markers of response prediction to immunotherapy in advanced NSCLC. This work may improve our understanding of tumor immunobiology and, if validated, offers several important opportunities to improve management and outcome of patients with advanced NSCLC and possibly also other tumors.

## Methods

### Patient enrolment

This study was approved by the Heidelberg University (S-296/2016, S-089/2019, S-916/2019) and Grosshansdorf Hospital ethics committee (AZ 12-238, AZ 19-286) and involved patients with advanced NSCLC treated with immunotherapy alone or immunotherapy in combination with chemotherapy, for which blood samples were available. The study performed in Grosshansdorf was registered in the German Clinical Trials Register (DRKS) under DRKS00018784 on 2019/11/18. The blood cell sorting study was registered under DRKS00022300 on 2020/06/29. The Thoraxklinik samples from Heidelberg originated from a biobank for which no DRKS registration was performed. All patients provided written informed consent. The Heidelberg patients were collected prospectively as published^45^ and provided by the Lungenbiobank Heidelberg and Biobank Nord for the present analysis according to the pertinent regulations. The Grosshansdorf patients were collected prospectively. Initially, a training cohort of 96 patients treated with PD-1 inhibitor monotherapy was assembled from Heidelberg, followed by an independent validation cohort of 99 additional patients who received PD-1 monotherapy in either Heidelberg (n=84) or Großhansdorf (n=15), as well as an independent control cohort of 139 patients who received chemoimmunotherapy in Heidelberg (Table 1). Diagnosis of NSCLC was performed in the Institute of Pathology Heidelberg and Borstel using tissue specimens according to the criteria of the current WHO classification (2015) for lung cancer, as described previously^46,47^. Clinical data and laboratory results were collected by a systematic review of patient records, as described^45^. The following clinical data were extracted: demographic, baseline clinical and tumor characteristics, including ECOG PS, smoking status, PD-L1 TPS, laboratory results, systemic and local anticancer treatments, date of progression, date of the last follow-up, and date of death. OS was calculated from start of immunotherapy to the time of last follow-up or death. For PD-L1 TPS assessment, the clones SP263 (Ventana/Roche, Mannheim, Germany) and E1L3N (Cell Signaling Technology Europe B.V.) were used in Heidelberg and Mannheim, respectively, and values trichotomized as < 1, 1-49, and ≥ 50%, reflecting the relevant thresholds in current treatment guidelines^48^.

### PAXgene collection and processing

PAXgene (PreAnalytiX, Hombrechtikon, Switzerland) blood samples were acquired as per manufacturer instructions, inverted immediately 10x and frozen at -20 °C within 2 hours. For long-term storage the tubes were transferred to -80 °C. *Training Cohort*. The PAXgene tubes were thawed overnight and extracted using the PAXgene Blood miRNA Kit (Qiagen, Venlo, Netherlands) as per manufacturer instructions. Eluates of 80 μl in RNase-free water were stored in -80°C. *Validation and Control Cohorts*. For the 223 Heidelberg patients, PAXgene tubes were thawed overnight, and RNA was extracted using PAXgene Blood RNA Kit (Qiagen, Venlo, Netherlands). The flow-through of the first column (RNA shredder column) was saved and stored in -20°C and used for further extraction of RNAs <200 nts of length. For this purpose, the alcohol content of the sample was brought to 50% (w/v) with isopropanol and the samples were processed further using PAXgene Blood miRNA Kit, starting from the RNA column step. The RNA was eluted in 40 μl RNase-free water and stored at -80°C. The RNA from PAXgene samples of the 15 Grosshansdorf patients was extracted using the QIAsymphony PAXgene Blood RNA Kit on the QIAsymphony SP liquid handling station (Qiagen, Venlo, Netherlands). The elution was performed at 72°C for 10 min in 200 μl of elution buffer. The RNA was aliquoted and stored at -80°C.

### Small RNA-seq

100 ng of total RNA, or 5 μl of total RNA (for samples with concentration lower than 20 ng/μl) were used for QIAseq^®^ miRNA Library Kit (Qiagen, Venlo, Netherlands). To block highly abundant miRNAs (miR-16-5p, miR-486-5p, miR-451a), custom blocking reagent FastSelect (Qiagen, Venlo, Netherlands) was used and added before the RT step. cDNA was amplified using primers (Supplementary Data 5) allowing unique dual indexing of the libraries and PCR products were cleaned-up using Mag-Bind TotalPure NGS beads (Omega Bio-Tek, Norcross, USA). PCR products following 18 cycles of amplification were assessed for size and uniformity using the Quantitative DNA kit on Fragment Analyzer (Agilent, Santa Clara, USA). The concentration of the PCR products was determined by Quant-iT dsDNA Assay-Kit (ThermoFisher Scientific, Waltham, USA), and equimolar library pools with up to 96 samples were prepared and sequenced on NextSeq 500 (Illumina, San Diego, California) with a 2.7 pM final pooled library concentration. A custom index 2 sequencing primer with the sequence 5’-GATCGTCGGACTGTAGAACTCTGAACGTGT-3’ was used.

### Blocking design and efficiency evaluation

We designed blocking oligonucleotides for the three most abundant miRNAs (hsa-miR-486-5p, hsa-miR-16-5p, hsa-miR-451a) within PAXgene whole blood libraries to target the nconsensus sequence defined from the alignment of the 20 most prevalent respective isomiR sequences (see Supplementary Fig. 1). Based on these consensus sequences, customized QIAseq® FastSelect RNA Removal Kits were obtained and deployed immediately prior to the reverse transcription step of library preparation (Fig. 1b). To evaluate the blocking efficiency, small RNA-Seq of the 96 PAXgene samples of the training cohort was performed using either an unblocked or blocked library preparation protocol (see section below). For illustration, the proportion of each miRNA in the total read counts was calculated for each sample. Additionally, the log2-transformed reads per million (RPM) value in each sample was calculated for the 1517 miRNAs that were detected in both libraries. The mean values of log2-transformed RPM values per miRNA in the unblocked and blocked samples were used for plotting as well as calculating the Pearson coefficient for unblocked and blocked samples.

### Statistical analysis

Non-penalized, univariable and multiviable Cox models for survival regression were fit in Python using the packages scikit-survival (version 0.15.1)^19^ and Lifelines (version 0.26.0)^49^ using the CoxPHSurvivalAnalysis and CoxPHFitter classes respectively with defult parameter settings. In order to overcome the p >> n problem while modelling survival in high dimensional data, a number of methods including discrete feature selection using univariable and stepwise selection have been proposed^50,51^. We have adapted a computational pipeline described by Shukla et al.^51^, using scikit-learn (version 0.24.2)^52^ and the SelectFpr (selecting miRNA features to which a Cox proportional hazards model could be fit with *P* < 0.05) and SequentialFeatureSelector (sequentially adding single miRNA features to a multivariable Cox proportional hazards model and assessing based on minimising the log-rank test *P* value between low/high risk cross-validated predictions) classes. Scikit-learn pipelines were used to couple the feature selection and training steps to create survival models and perform inference on new data.

Random survival forests from the scikit-survival package were used to train forests using the parameter settings: n_estimators=1000, min_samples_split=10, min_samples_leaf=15, max_features=“sqrt”. Feature importance was assessed using the permutation based method from the package ELI5 (version 0.11.0) by estimating the reduction in model concordance index when removing the association between survival and each feature in turn (through random shuffling).

Patients were stratified into high/low risk groups by the median risk score in the training data, based on the cutpoint definition of similar previously described prognostic gene expression signatures^53–55^. Prognostic performance of the miRisk score was assessed in a multivariable Cox proportional hazards model with additional clinicopathological covariates (PD-L1, ECOG, gender, age, therapy line, substance, histology, smoking status, ANC, ALC). The survival function was estimated using the Kaplan–Meier method, and groups were compared with the log-rank test.

Time dependent ROC curve analysis was performed in R with the package timeROC and bootsrapping was performed to quantify differences in AUC between models (e.g., miRisk versus miRisk + PD-L1).

### qRT-PCR

Catalogue and custom miRCury LNA miRNA PCR assays were ordered from Qiagen (Venlo, Netherlands) for biomarker miRNAs and housekeeping miRNAs (Supplementary Data 6). 500 ng RNA was used for cDNA synthesis, or when the concentration was insufficient, up to 6.5 μl of RNA volume was used. cDNA was then diluted 1:80, 10μl total volume reaction was set up according to manufacturer’s instructions, and cycled according to the following protocol: 2 min at 95°C, 40x 10s at 95°C 60s at 56°C in AB Flex 6 cycler in 384-well format. A melt curve analysis was performed at 60-95°C. The C_†_ values were determined and exported using QuantStudio RealTime PCR Software version 1.3. For the training cohort of 95 patients, the C_t_ values of the five miRisk miRNAs were determined in triplicates. ΔC_t_ values were calculated by subtracting the mean of the C_t_ values of “housekeeping” miRNAs that were previously defined by NGS on the basis of low variance. Based on these ΔC_t_ values the fold change (2^ΔΔCt^) and t-test between samples from patients with low or high risk according to the miRisk score was calculated per signature miRNA.

### Purification of blood cells

Cell sorting was performed immediately after donation of human whole blood. An overview is provided in Supplementary Fig. 4. Whole blood was collected in S-Monovette^®^ EDTA K3 tubes (Sarstedt AG & Co. KG, Nümbrecht, Germany). To isolate CD4+ T-cells, CD8+ T-cells, monocytes, and B-cells directly from whole blood, 350 μL MicroBeads of the corresponding kit (see Supplementary Data 7) were added separately to 7 mL EDTA K3 blood. After incubating for 15 min at 4° C, cell suspensions were washed by adding isolation buffer to the cell suspension, performing a centrifugation step of 5 min at 400 x *g* and 4° C, and discarding the supernatant. Next, the resulting pellets were resuspended in 1 mL isolation buffer. Lastly, a positive selection of the magnetic labelled cells was performed with the autoMACS® Pro Separator (Miltenyi Biotec GmbH, Bergisch Gladbach, Germany). For the isolation of neutrophils and NK cells, the corresponding MACSxpress^®^ Whole Blood Isolation reagent (Supplementary Data 7) was added in a ratio of 1:2 to the whole blood. Next, tubes were rotated in a MACSmix^™^ Tube Rotator (Miltenyi Biotec GmbH, Bergisch Gladbach, Germany) for 5 min at room temperature. After 15 min on a MACSxpress Separator (Miltenyi Biotec GmbH, Bergisch Gladbach, Germany) the supernatant containing the cell populations was collected. To further purify neutrophils and NK cells, a lysis of erythrocytes was performed by adding 20 mL 0.2% NaCl solution (Merck KGaA, Darmstadt, Germany) for 20 s and 20 mL 1.6% NaCl solution thereafter and the cell pellet was saved after centrifugation for 5 min at 300 x *g* and 4 °C. To isolate human thrombocytes, basophils, and eosinophils, whole blood was diluted in a ratio of 2:3 with PBS (Thermo Fisher Scientific, Waltham, USA) and layered over the density gradient medium Histopaque^®^-1077 (Merck KGaA, Darmstadt, Germany) in a ratio of 3:5. After a centrifugation for 20 min at 600 x *g* and RT (no brake), the different layers were isolated immediately.

The uppermost plasma layer was used to isolate platelets. After an additional centrifugation step for 15 min at 500 x *g* and RT, the supernatant was discarded, and the pellet was resuspended in 600 μL isolation buffer and 150 μL CD61 beads (Supplementary Data 7) were added for positive selection. After an incubation time of 15 min at 4 °C, the pellet was washed, resuspended in 500 μL isolation buffer (0,5% albumin, 2 mM EDTA in PBS) and thrombocytes isolated with the autoMACS Pro Separator. The interphase ring layer was washed twice with isolation buffer. The isolation of basophils was performed in a two-step procedure with the Diamond Basophil Isolation Kit (Supplementary Data 7). First, the cell pellet was resuspended in 300 μL isolation buffer per 10^8^ cells, 100 μL FcR Blocking Reagent per 10^8^ cells, and 100 μL Basophil Biotin-Antibody Cocktail per 10^8^ cells. After incubating for 10 min at 4 °C, 300 μL isolation buffer per 10^8^ cells and 200 μL Anti-Biotin MicroBeads per 10^8^ cells were added. After incubating again for 10 min at 4 °C, cells were washed, resuspended in 500 μL isolation buffer and negative selection was performed with the autoMACS Pro Separator. Afterwards, the enriched fraction was washed, resuspended directly in 100 μL CD123 MicroBeads and incubated for 15 min at 4 °C. After washing the resulting cell pellet was resuspended in 500 μL isolation buffer and used for positive selection on autoMACS Pro Separator.

The bottom layer was purified by performing several erythrocyte lysis steps. Next, the cell suspension was centrifugated for 5 min at 300 x *g* and 4 °C. The pellet was then resuspended in 40 μL isolation buffer per 10^7^ cells and in 10 μL Eosinophil Biotin-Antibody Cocktail per 10^7^ cells (Supplementary Data 7). After an incubation time of 10 min at 4 °C, 30 μL isolation buffer per 10^7^ cells and 20 μL Anti-Biotin MicroBeads per 10^7^ cells were added to the cell suspension, incubated 15 min at 4 °C, washed and finally resuspended in 500 μL isolation buffer. Eosinophils were isolated with the autoMACS Pro Separator via a negative selection.

To separate erythrocytes from human whole blood, 5 mL whole blood was centrifuged for min at 2500 x *g* and RT. 1 ml plasma was centrifuged for 2 min at 13,000 x g, aliquoted and stored in -80°C. The remaining pellet was resuspended in isolation buffer, filtered with a 40 μm cell strainer (Greiner Bio-One GmbH, Frickenhausen, Germany) and diluted in a ratio of 1:3 with isolation buffer. Typically, 3×10^7^ erythrocytes were centrifuged for 10 min at 300 x *g* and 4 °C, cell pellet was resuspended in 240 μL isolation buffer and 60 μL CD235a MicroBeads (Supplementary Data 7) and incubated for 15 min at 4 °C. After washing, the cell pellet was resuspended in 500 μL isolation buffer and used for positive selection on autoMACS Pro Separator.

The yield of all purified cell populations was determined, and cell pellets were directly lysed in Qiazol and stored at -80°C.

### RNA isolation of sorted cell pellets

Cell lysates in Qiazol were thawed on ice and used for RNA isolation using miRNeasy mini kit combined with minElute columns. The RNA was eluted in 18 μl of water and stored at - 13 80°C.

### Clinical differential and epigenetic PCR blood counts of 12 healthy donors

Absolute erythrocyte and platelet counts were obtained from clinical differential blood counts from the University of Heidelberg. Absolute quantifications (cells/ml) of neutrophils, basophils, eosinophils, monocytes, NK cells, CD4+, CD8+, CD19+ cells were performed from EDTA whole blood by Epiontis, Precision for Medicine using epigenetic PCR^56^.

### Flow cytometry quality control of sorted cells

Typically, 0.5×105 cells were diluted in 2 mL isolation buffer and centrifuged afterwards for 5 min at 300 x g and RT. The cell pellets were resuspended in 100 μL isolation buffer, 5 μL of Human TruStain FcX™ (BioLegend; San Diego, USA) was added and pre-incubated for 10 min at RT. Afterwards, the corresponding antibodies were added (Supplementary Data 8) and incubated for 20 min at 4 °C in the dark. Next, 2 mL isolation buffer was added, centrifuged for 5 min at 300 x g and RT and resuspended in 300 μL isolation buffer. The FACS measurements were performed on a BD FACSVerse™ flow cytometer (BD Biosciences, Franklin Lakes, USA) and analysed using BD FACSuiteTM software v1.0.5 (BD Biosciences, Franklin Lakes, USA). The full gating strategy is provided Supplementary Figure 8.

### Sorted blood cell NGS data processing

Raw sequences were adapter trimmed and depleted of PCR duplicates based on unique molecular identifiers (UMIs) using a customized R script. To annotate the small RNA sequences the collapsed reads were mapped with Bowtie (version 1.2.3)^57^ to the miRNA hairpin sequences obtained from the miRbase database (version 22)^58^. The counts of canonical miRNA and isomiR sequences were summed up to generate count matrices for the detected miRNAs.

### Deconvolution of PAXgene RNA profiles into cellular origins

We computed the relative contribution of each of the 10 analyzed cell types to the expression of an individual miRNA in whole blood. For this purpose, the RPM values of miRNA *m* in cell type *c* and patient *p*, (*x*_*m,c,p*_) are multiplied with a scaling factor (α_*c,p*_). This scaling factor accounts for the small RNA content per sorted cell and the number of cells of type *c* that are present in the blood of patient *p* (n_blood_count_). The small RNA content of each sample is calculated as the product of the small RNA concentration (c_sRNA_) and the elution volume of the sorted cells (V_elution_) divided by the number of sorted cells (n_sorted_count_):

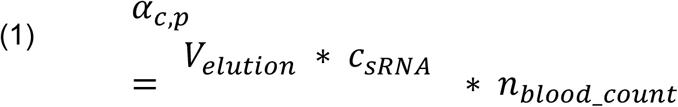

The mean of the scaled RPM values of miRNA *m*, per cell type *c*, is aggregated across all patients, *P*:

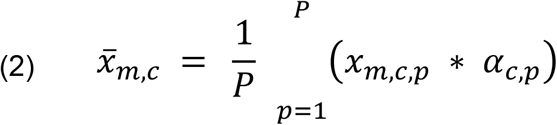

Finally, the proportion of the scaled mean expression values of each cell type *c* was calculated per miRNA *m*:

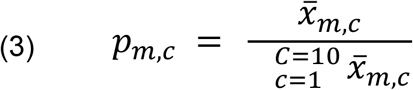

These proportions were then used to visualize the cell type distribution of miRNAs of interest and to generate a clustermap of all miRNAs that have read counts > 0 in whole blood PAXgene small RNA sequencing libraries, using the Python library seaborn (version 3.8)^59^.

### Correlation miRNA expression and cell type blood count

To investigate whether the expression of the 44 survival-associated miRNAs is correlated with the abundance of the cell type they are predominantly expressed in, we calculated Pearson coefficients for RPM values of each miRNA and the blood counts of each cell type based on the training cohort (n=95). The sample with the ID 464_0033 was excluded from this analysis as it had an unusually high monocyte count (5.2 standard deviations above the mean).

### Target Prediction and Pathway Analysis

Predicted target mRNAs of the 44 survival-associated miRNAs were defined by the presence of conserved and non-conserved miRNA response elements (MREs) within their 3’ UTR using TargetScan (version 7.2,)^24^. KEGG Enrichment Analysis of the predicted targets was performed using the R package ClusterProfiler (version 3.18.1)^60^.

## Data Availability

Data is available from the corresponding author upon reasonable request.

## Data Availability

Anonymized small RNA sequencing data are available with controlled access approval through the European Nucleotide Archive under accession number PRJEB50502. All other data supporting the findings of this study are available from the corresponding author on reasonable request.

## Code Availability

Data analysis was performed in Python (3.8.8) with the packages scikit-learn (version 0.24.2), scikit-survival (version 0.15.1), and Lifelines (version 0.26.0), and RStudio (Version 1.3.1073) with the package timeROC (version 0.4), and described in the methods. Raw code will be made available from the corresponding author upon reasonable request.

## Acknowledgements

We thank Stephanie Axenfeld, Urszula Jagos-Ihde, Claudia Genest, Marlen Szewczyk, Milena Schmidt and all other contributing medical personnel at the LungenClinic in Großhansdorf for their outstanding work with patient enrollment and data collection. We also thank Elena Gleim, Elena Neumüller at Hummingbird Diagnostics GmbH and Helena Schock, PhD for their excellent technical assistance during the study conduct. We thank Mihaela Zavolan and Anastasiya Börsch for discussions of the cell sorting analyses. Finally, we thank all study participants and their families.

## Author Contributions

T.R., P.C., B.R.S. designed and planned the study. P.C., F.H., K.T., S.H., T.M., S.U., M.A.W., M.E., F.L., H.S., K.F.R., M.R., M.T., organized clinical enrolment and sample and data collection. R.H., J.S., O.P., J.F., J.Sk., D.N., A.D.M., J.C., F.U., Q.W. processed biological material and performed experiments. T.R., J.Je., M.K., P.K., M.H., T.S., N.M., Q.F., Q.W., T.S.S., P.C., B.R.S. analysed and interpreted the data. T.R., P.C., B.R.S. wrote the manuscript. All authors approved the final version.

## Competing Interests

P.C. declares research funding from AstraZeneca, Novartis, Roche, Takeda, and advisory board/lecture/educational fees from AstraZeneca, Boehringer Ingelheim, Chugai, Kite, Novartis, Pfizer, Roche, and Takeda. M.R. reports receiving honoraria for lectures and consultancy from AstraZeneca, Amgen, BMS, Boehringer-Ingelheim, Lilly, Merck, MSD, Mirati, Novartis, Sanofi, Pfizer, Roche. K.F.R. reports payments or honoraria from Boehringer Ingelheim and Astra Zeneca, Novartis, Roche, Chiesi Pharmaceuticals, Regeneron, Sanofi and Berlin Chemie outside the submitted work. M. T. discloses honoraria from AstraZeneca, Bristol-Myers Squibb, Boehringer Ingelheim, Celgene, Chugai, Lilly, MSD, Novartis, Pfizer, Roche, Takeda, Sanofi, Beigene, GSK and research funding from AstraZeneca, Bristol-Myers Squibb, Roche and Takeda. T.R., R.H., J.Je., S.H., P.K., M.K., M.H., T.S. J.F., J.Sk., D.N., A.D.M, J.C., B.R.S. are employees of Hummingbird Diagnostics and hold company stock options. T.R., R.H., J.Je., T.S., B.R.S. are inventors of patent applications related to response prediction fo immunotherapy submitted by Hummingbird Diagnostics. T.S.S. serves on the scientific advisory board of Hummingbird Diagnostics. N.M. received consulting fees from Hummingbird Diagnostics. The remaining authors declare no competing interests.

## Notes

### Funding Statement

This study did not receive any funding.

### Author Declarations

This study was approved by the Heidelberg University (S-296/2016, S-089/2019) and Grosshansdorf Hospital ethics committee (AZ 12-238).

