## Supplementary Figures for "A blood-based miRNA signature with prognostic value for overall survival in advanced stage non-small cell lung cancer treated with immunotherapy" (from another version of this preprint: version 1, 2021-11-01)

### Supplementary Figure 1

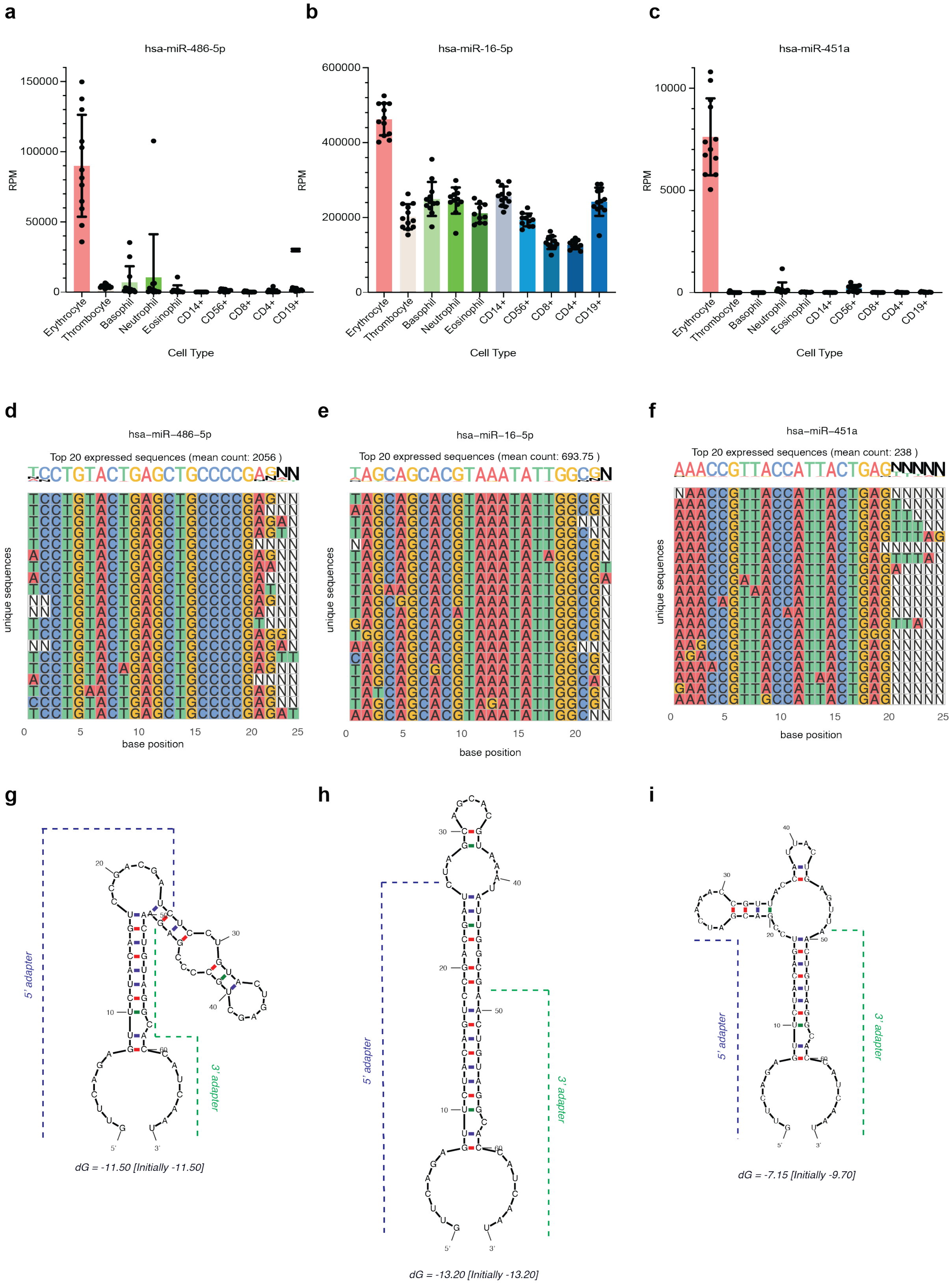

**Supplementary Figure 1 | Design of antisense LNA blocking oligonucleotides. a-c** Expression of three highly abundant miRNAs in the main cell types of peripheral blood. hsa-miR-486-5p and hsa-miR-451a exhibit almost solely erythroid origin. hsa-miR-16-5p is highly expressed in all blood cell types. **d-f** For each of the intended miRNA targets, motif analysis was performed on the 10 most abundant isomiRs to identify the minimal consensus sequence against which the blocking oligonucleotides were designed. **g-i** The structure of adapter ligated miRNAs was predicted *in silico* using RNAfold. hsa-miR-16-5p (h) exhibits an extremely stable stem loop making it a probably preferred substrate for the T4 RNA ligase. This likely explains the dominant abundance of hsa-miR-16-5p in the library (“jackpotting”).

### Supplementary Figure 2

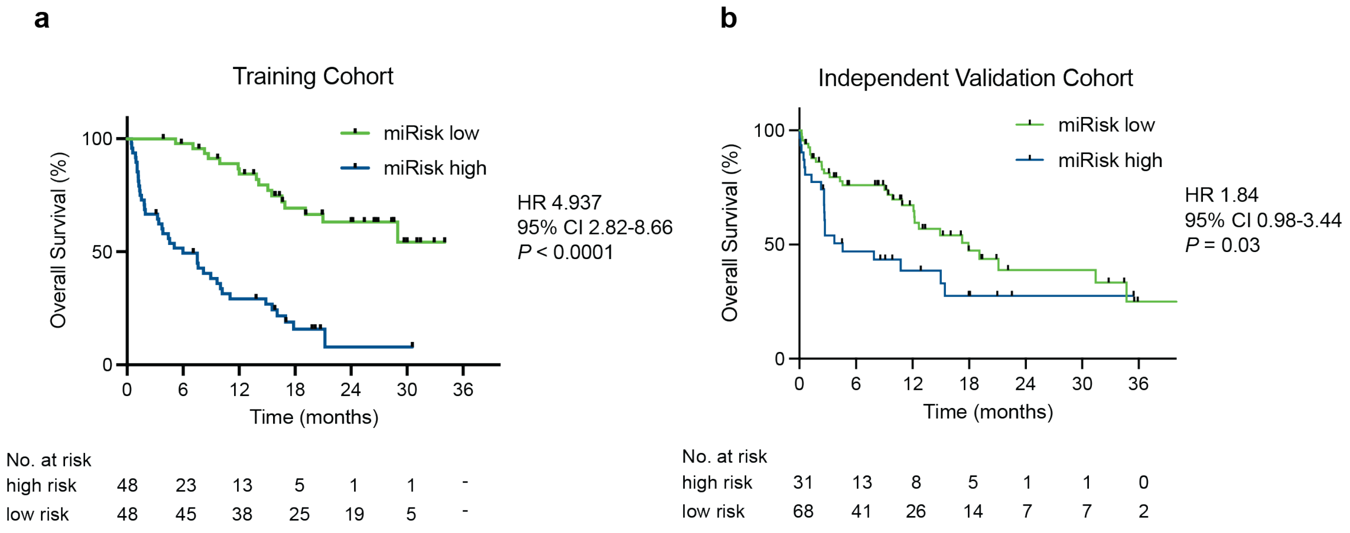


**c**

| Weight | Feature |
|---|---|
| **0.0108 ± 0.0179** | **hsa-miR-2115-3p** |
| 0.0046 ± 0.0049 | hsa-miR-150-5p |
| 0.0038 ± 0.0035 | hsa-miR-2115-5p |
| 0.0037 ± 0.0031 | hsa-miR-155-5p |
| 0.0035 ± 0.0030 | hsa-miR-143-3p |
| **0.0034 ± 0.0024** | **hsa-miR-224-5p** |
| 0.0032 ± 0.0039 | hsa-miR-5001-3p |
| **0.0029 ± 0.0037** | **hsa-miR-218-5p** |
| 0.0027 ± 0.0020 | hsa-miR-338-3p |
| 0.0024 ± 0.0013 | hsa-miR-219b-5p |
| 0.0022 ± 0.0029 | hsa-miR-1180-3p |
| 0.0022 ± 0.0034 | hsa-miR-3667-3p |
| 0.0021 ± 0.0019 | hsa-miR-10399-5p |
| 0.0021 ± 0.0010 | hsa-miR-31-5p |
| 0.0020 ± 0.0011 | hsa-let-7f-5p |
| 0.0019 ± 0.0020 | hsa-miR-126-3p |
| 0.0018 ± 0.0027 | hsa-miR-1273h-3p |
| 0.0018 ± 0.0015 | hsa-miR-16-1-3p |
| 0.0017 ± 0.0030 | hsa-miR-7706 |
| 0.0017 ± 0.0032 | hsa-miR-30a-5p |

**Supplementary Figure 2 | Overall survival of low and high-risk patients stratified by random forest model. a-b** Comparison of OS between low-risk and high-risk patients in the training cohort (n = 96), and independent validation cohort (n=99). **c** Random forest feature importance as assessed by permutation testing. Weights indicate the decrease in concordance index attributed to each feature. Bold miRNAs are shared with the miRisk signature.

### Supplementary Figure 3

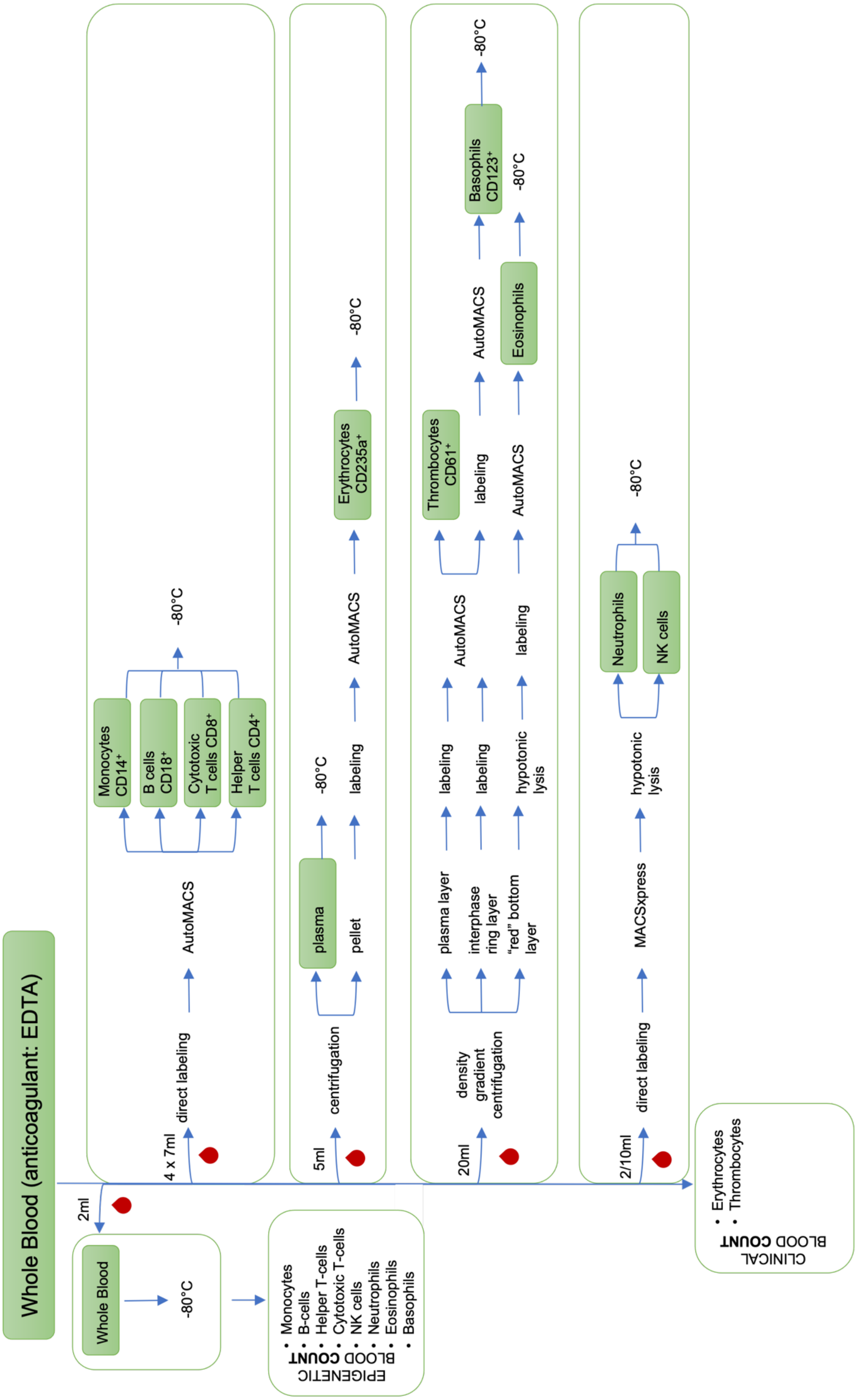


**Supplementary Figure 3 | Blood cell sorting scheme** Peripheral blood cells were sorted and enumerated according to the flow chart.

### Supplementary Figure 4

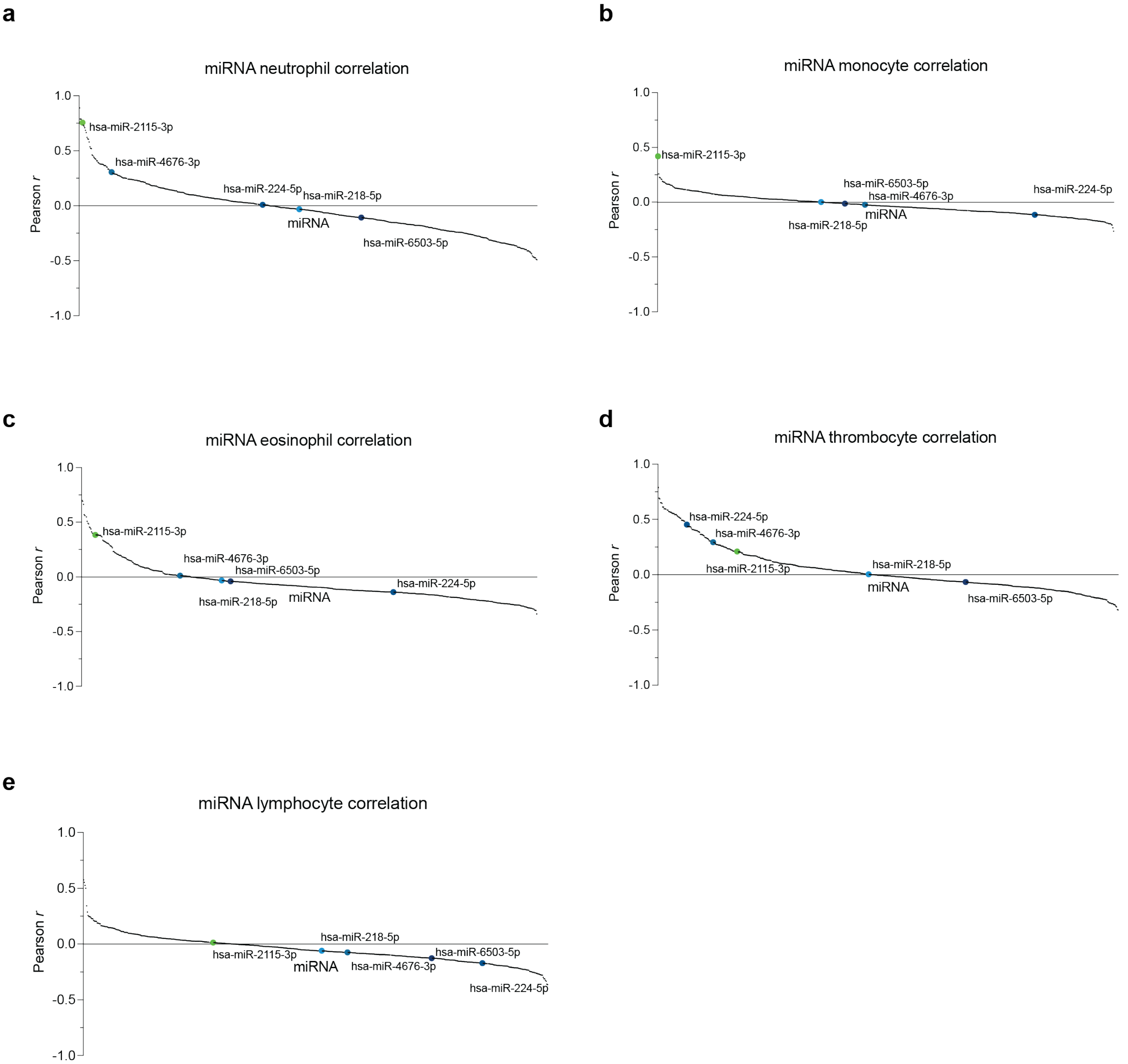


**Supplementary Figure 4 | Correlation analysis of miRNA expression in RPM to clinical differential blood count. a-e** Ranked Pearson correlation coefficient r between miRNA expression and blood count for 5 cell types, calculated in the training cohort (n=96). The 5 miRisk miRNAs are highlighted. A correlation is observed between miR-2115-3p and its predominant cell types of origin (neutrophils and monocytes). In contrast, miR-6503-5p demonstrates no correlation to its predominant cell type of origin (monocytes).

### Supplementary Figure 5

**a**

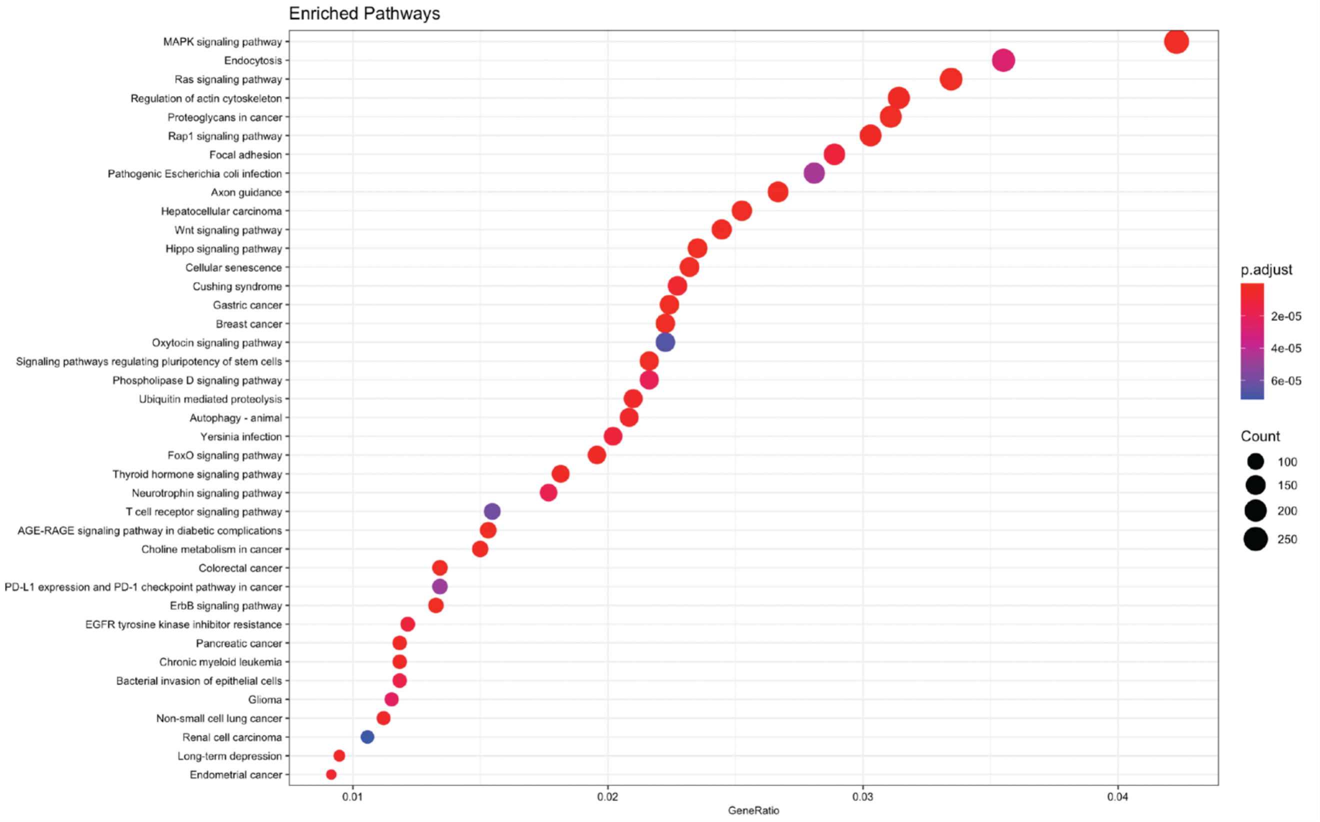


**b**

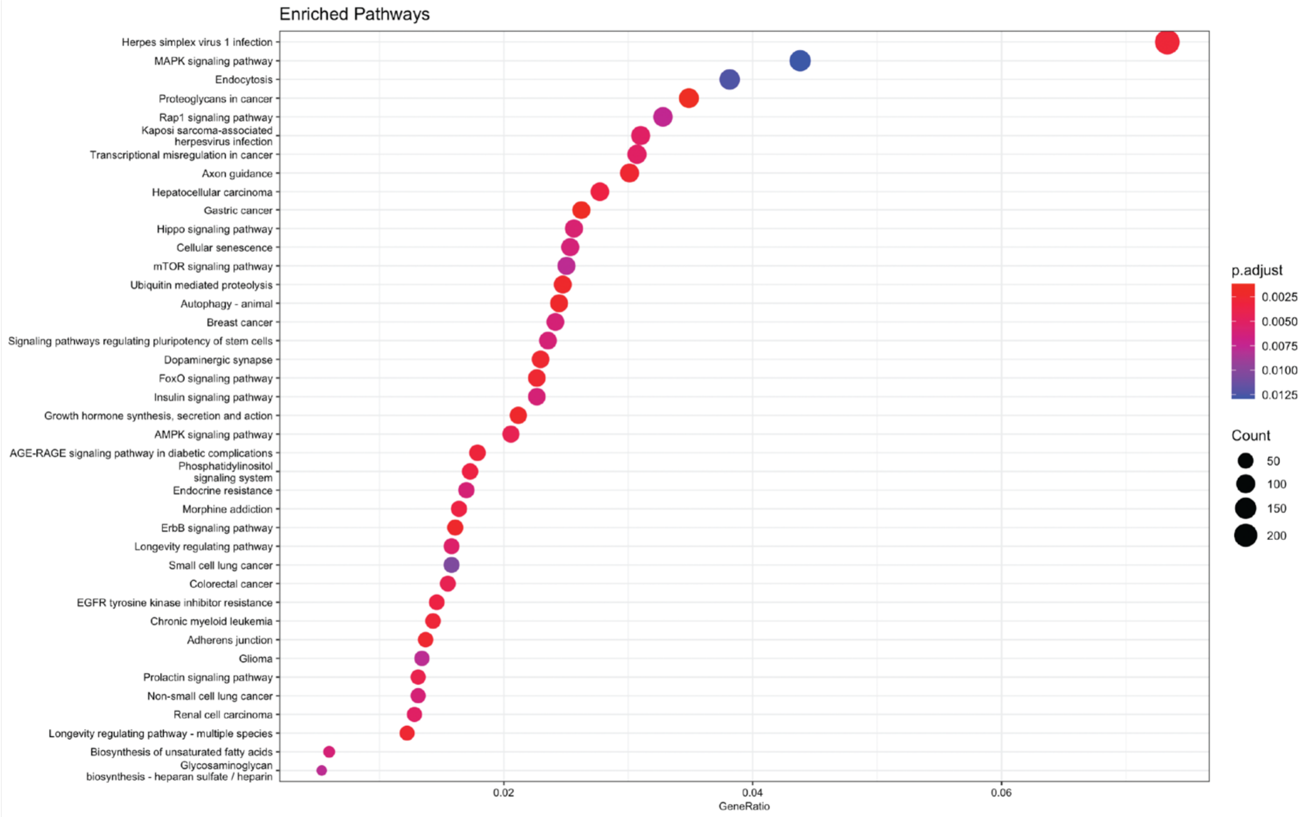

**Supplementary Figure 5 | Pathway enrichment analysis of predicted gene targets. a** Enriched pathways for targets of the 44 survival-associated miRNAs. Of the 6337 predicted targets listed in the KEGG database, 268 are part of the MAPK signaling pathway (1.16-fold enrichment compared to a 294/8095 background ratio, adjusted P-value = $1.58x10^{-7}$) and 85 are part of the PD-L1/PD-1 checkpoint pathway (1.22-fold enrichment compared to a 89/8095 background ratio, adjusted P-value = $4.84x10^{-5}$). **b** Enriched pathways for targets of the five miRisk miRNAs. With 147 of 3355 predicted targets listed in the KEGG database, the MAPK signaling pathway is the second most significantly enriched pathway (1.21-fold enrichment compared to a 294/8095 background ratio, adjusted P-value = $1.29x10^{-2}$). Pathway overrepresentation compared to the expectation was tested by hypergeometric testing. *P* values were corrected for false discovery using the Benjamini-Hochberg method. The plotted gene ratio is the proportion of predicted target genes that map to a certain pathway. The dot size is dependent on the number of mapped genes per pathway. The color of the dots reflects the adjusted *P* value level of the enrichment testing.

### Supplementary Figure 6

**a**

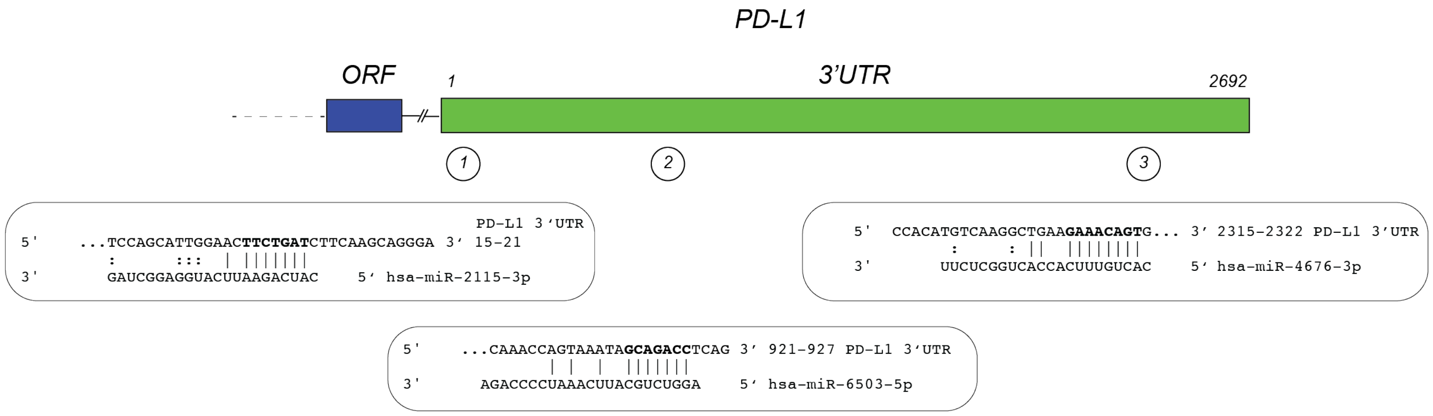


**b**

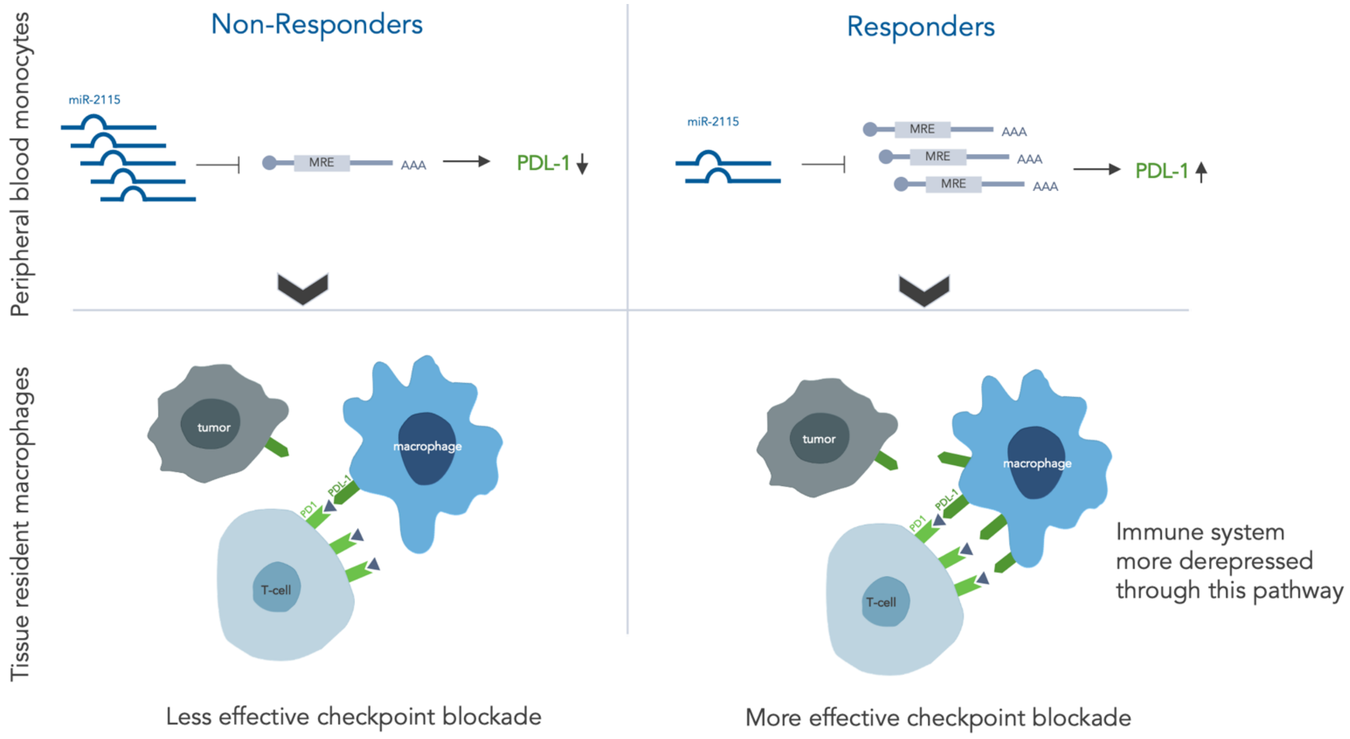


**Supplementary Figure 6 | Direct interaction between miRisk miRNAs and PD-L1 and the proposed mechanism of action. a** Schematic representation of the 3' untranslated region (UTR) of PD-L1 indicating the location of predicted miRisk miRNA target sites and base pairing between miRNA seed and target site. **b** Potential mechanism of action is that lower expression of miRisk miRNAs in peripheral myeloid cells of responders leads to reduced expression of PD-L1, which is maintained upon migration into the TME. This creates a TME in which abundant PD-L1 expression promotes immunosuppression but generates susceptibility to immune checkpoint blockade.
